# Face-to-face interventions to encourage enrolment in cardiac rehabilitation; a scoping review protocol

**DOI:** 10.1101/2021.02.19.21252058

**Authors:** Birgit Rasmussen, Sasja Jul Håkonsen, Bente Skovsby Toft

## Abstract

**Introduction:** Cardiac rehabilitation has become an integral part of secondary treatment of cardiovascular heart disease. Despite evidence demonstrating that cardiac rehabilitation improves prognoses, reduces disease progression, and helps patients to find a new foothold in life, many patients do not enrol. Face-to-face interventions can encourage patients to enrol, however, it is unclear which strategies have been developed, how they are structured in a hospital context, and whether they target the life-world of the patients. The objective of this scoping review is to map and evaluate the nature and characteristics of studies that have reported on face-to-face interventions to encourage patients to enrol in cardiac rehabilitation.

**Methods and analysis:** This review will be guided by the Joanna Briggs Institute Methodology for Scoping Reviews. A search strategy developed in cooperation with a research secretary will be applied in six databases including studies published from 2000 in English, Danish, Norwegian, Swedish, and German with no restriction on publication type or study design. Studies involving adult patients with ischemic heart disease or heart failure will be included. Studies providing the intervention after enrolment in cardiac rehabilitation will be excluded. Study selection will be performed independently by two reviewers. Data will be extracted by two reviewers using predefined data charting forms. The presentation of data will be a narrative summary of the characteristics and key findings to facilitate the integration of diverse evidence, and as we deem appropriate will be supported by a diagrammatic or tabular presentation.

**Ethics and dissemination:** This scoping review will use data from existing publications and does not require ethical approval. Results will be reported through publication in a scientific journal and presented on relevant conferences and disseminated as part of future workshops with professionals involved in communication with patients about enrolment in cardiac rehabilitation.

**ARTICLE SUMMARY:** *Strength and Limitations of this study:*

- This protocol outlines a rigorous design that includes the use of an established scoping review methodology and a search strategy developed in cooperation with a research librarian
- The search strategy has no restrictions to study design, includes five different languages, and will cover six different databases
- As this review is inclusive to all study types and aims to provide an overview of the landscape of interventions to encourage enrolment in cardiac rehabilitation a quality assessment will not be performed
- While the review will be non-discriminant towards article study types and methodologies, a limitation of the study is that books and grey literature will not be included

## INTRODUCTION

Cardiovascular disease is among the leading causes of mortality and disability worldwide. Each year ischemic heart disease gives rise to an estimated 8,92 million deaths globally[1] and the estimated one-year mortality in people living with heart failure is 20-30%.[2] People living with ischemic heart disease and heart failure are likely to experience diminished quality of life, readmissions, and debilitating symptoms.[3,4] Cardiac rehabilitation (CR) is evidenced to reduce mortality, reduce the risk of readmission, improve quality of life[5,6] and it can reduce anxiety and depression; conditions found to worsen the prognosis for survival after cardiac events.[7] As such since the beginning of the millennium participation CR has been acknowledged as a crucial therapeutic tool.[8] However, despite significant improvement in prognoses and personal benefits from participating in CR, many patients do not enrol.[9]

The treatment of ischemic heart disease and heart failure has been significantly improved over the last decades and aims to limit disease progression, prevent or reduce complications, and to eliminate ischemic symptoms. The primary treatment is medical therapy, device therapy, and surgical revascularization, whereas CR is becoming an integral part of standard secondary treatment targeting risk modification to promote recovery and prevent further cardiac events.[10,11] CR programs include education, counselling, and behavioral strategies to improve health behavior in relation to nutrition, smoking, stress, and training, and is offered to assist patients with heart disease to move forward and live healthy and satisfying lives.[12] Participating in CR helps patients find a new foothold in life.[13] Attendees feel supported by being with peers during the course of the rehabilitation program and from the knowledge and encouragement they receive from the CR staff. [14] However, though the benefits from participation in CR are manifest, less than half of patients participate[15–17] and there is a need to consider strategies to encourage more patients to enrol.

Barriers for participation in CR seem to be multiple and complex and appear across factors such as age, social position, and culture. Particularly women, people older than 70 years, younger people, smokers, people with more comorbidity, people with reduced functioning, people living alone, unemployed people, and people who have a lower income are more likely to abstain from participation.[18] Contextual factors such as distance, transportation difficulties, family commitment and opinions of significant others are reported to be barriers for enrolling in CR.[19]. Though systematized referral is found to be a key to secure that patients enrol in CR,[18] many patients abstain from participation even when systematized referral is implemented in hospital practices.[9] The combination of systematized referral and discussions between the individual patient and a healthcare professional, the so called ‵liaison’ strategies, have been found more effective than systematized referral on its own, and the incorporation of these interventions into standard, in-hospital pathways has been recommended.[20] Particularly liaison strategies involving at least some elements of face-to-face contact and when delivered by healthcare professionals have proven to promote enrolment.[21]

Studies on face-to-face interventions are limited[21] and the strategies used to influence enrolment vary, e.g. the provision of education and advice about CR,[22] social support,[23,24] the targeting of patients’ intention to enrol,[25] their illness perception,[26] self-efficacy,[23] and beliefs about CR,[27,28] or the focusing on individual barriers for enrolment. [25] Some studies proven to be effective in increasing enrolment rates use psychologically based behavioral theories to guide the development of the intervention i.e., Banduras theory of self-efficacy,[23] Leventhal’s self-regulation theory,[26] and theory of planned behavior.[25] However, interventions targeting patients’ behavior, intentions to attend, and health belief may fail to convey how CR can be connected with their everyday life.[29] The experience of health in a person’s life goes beyond the modification of risks and intentions to live a life without disease,[30] and when patients make decisions regarding their participation in CR, existential thoughts concerning their own well-being and suffering can be essential.[31] Hence, it seems pivotal that development of interventions take into account a person’s life-world and contemplate both patients’ agency and vulnerability.[32]

For the patients participation may conflict with their emotions, beliefs or sense of identity[29] and thoughts about one’s own mortality, and a wish to get back to normal and avoid worries have proven to be a barrier for participation in CR. Additional barriers are documented to be patients’ experiences of the nature and suitability of the program; accessibility; negative experiences of the healthcare professionals providing the services; and the view that CR is beneficial for others than one self, e.g. those who are more fit or more ill.[14] Though these barriers perceived by patients are acknowledged to influence their decision to enrol,[14] they rarely are targeted,[33] and a person-oriented approach where patients’ experiences and knowledge are taken into account is called for when barriers for enrolment are addressed.[34]

In-hospital face-to-face strategies to encourage participation in CR and bridge the gap between referral and enrolment are emerging. However, not much is known about the current scope of face-to-face interventions developed to facilitate enrolment in CR, their characteristics and content, how they are structured in a hospital context, and whether they are effective. A preliminary search of PUBMED, the Cochrane Database of Systematic Reviews and the *JBI Evidence Synthesis* identified a systematic review updated in 2019 on interventions to promote utilization of CR. However, the study only included randomized controlled trials to assess the effectiveness of interventions to increase utilization and only briefly considered the characteristics of the studies.{Santiago de Araujo Pio, 2019 #1328} No current or underway scoping reviews or systematic reviews on the topic of this scoping review were identified.

There is a need to gather the knowledge and experience from existing intervention studies to improve and develop interventions including how to support a person-oriented approach. The mapping and evaluation of evidence on face-to-face interventions encouraging patients to enrol in CR will provide an overview and a greater understanding of the use and characteristics of existing interventions and/or identify gaps of importance for future research. Also, an understanding of the outcomes evaluated in the current literature will aid clinicians in understanding the applicability of face-to-face interventions. With this scoping review we aim to map and evaluate the nature and characteristics as well as the outcomes evaluated of studies that have reported on face-to-face interventions to encourage enrolment in CR.

## METHODS AND ANALYSIS

The proposed scoping review will be conducted in accordance with the JBI methodology for scoping reviews[35] and will be reported according to the PRISMA Extension for Scoping Reviews.{Tricco, 2018 #1422} We follow the JBI reviewer’s manual to secure a contemporary methodological framework promoting clarity and rigor of the review process and to facilitate knowledge transfer to research and practice. The steps outlined in the JBI reviewer’s manual to be used are: (1) identifying the research question; (2) developing the inclusion criteria; (3) defining the search strategy; (3) study selection; (4) data extraction; and (5) presentation of the results.

### Review questions

The primary objective of this scoping review is to map and evaluate the nature and characteristics of studies that have reported on face-to-face interventions to encourage enrolment in CR. However, the review also will provide a narrative view on the extent of the evaluations of the interventions and a descriptive review of the effectiveness. The scoping review will map evidence pertaining to the following research questions:

1. What is the extent, range and nature of literature on face-to-face interventions to encourage enrolment in CR for adults with ischemic heart disease and heart failure?
2. What are the characteristics of the interventions?
  a. What factors considered to influence enrolment are targeted?
  b. What patient-experienced barriers and facilitators known to influence enrolment are targeted?
  c. How are patients’ experiences and knowledge taken into account?
  d. What outcomes are evaluated?
  e. What similarities and/or differences across the interventions exist?

The primary focus of this review is to make an account of existing interventions and their contents. Both experimental, quantitative, and qualitative study designs can provide evidence relevant to the objective of this study design.

### Inclusion criteria

#### Participants

This review will consider studies that include adults (older than 18 years). We include studies with participants with heart failure, participants who have had a myocardial infarction, have undergone surgical revascularization (coronary artery bypass grafting, percutaneous coronary intervention), or who have angina pectoris or ischemic heart disease defined by angiography, and who after surgery or medical treatment will be or have been offered a CR referral. Within these studies, information about intervention characteristics, participants’ experiences, and barriers for enrolment will be included. Studies focusing on participants that have already enrolled in a CR program will be excluded. We will exclude studies which only include participants with atrial fibrillation or with heart transplants, implanted with cardiac-resynchronization therapy or defibrillators, or who have had heart valve surgery.

#### Concept

This review will consider studies that investigate or explore face-to-face interventions including online face-to-face interventions performed by healthcare providers to encourage enrolment in CR. At least some part of the intervention should be face-to-face but can also take place in combination with other follow-up strategies i.e., telephone calls. This review will consider studies that are based in a hospital setting in any geographical area. Studies only providing the intervention outside the hospital, i.e., providing home visits, will be excluded. In-hospital includes telephone calls or other contacts from hospital healthcare providers to patients as a follow-up on a face-to-face intervention.

#### Context

CR is defined as a supervised or unsupervised inpatient, outpatient, community-or home-based intervention which includes some form of exercise training that is applied to a cardiac patient population. CR could be exercise training alone or exercise training in addition to psychosocial or educational interventions, or both (i.e., “comprehensive CR”).

#### Types of sources

This scoping review will consider quantitative, qualitative and mixed methods study designs for inclusion. In addition, systematic reviews will be considered for inclusion in the proposed scoping review. Text (for example political documents or government recommendation) and opinion papers will be excluded. Articles published in English, Danish, Norwegian, Swedish, and German will be included. Only studies published in 2000 or later will be included.

### Search strategy

The search strategy will aim to locate both published primary studies and reviews in databases with peer-reviewed literature. An initial limited search of PubMed and CINAHL (EBSCO) will be undertaken to identify articles on the topic. The text words contained in the titles and abstracts of relevant articles, and the index terms used to describe the articles will be used to develop a full search strategy for the databases PubMed, CINAHL, Cochrane Central Register of Controlled Trials, Embase, PsycINFO, and PEDro. The research strategy will be developed and refined in cooperation with a research librarian and will be adapted for each included information source. The preliminary search strategy for PubMed is presented in Appendix I. Only English search terms will be used. The reference lists of articles included in the review will be screened for additional papers. Authors of included studies will be contacted if further information about the study is required. A complete search strategy for CINAHL is presented in Table I.

**Table I:** search strategy for CINAHL. CINAHL (EBSCO host). Search conducted on February 2021

| Search | Query | Records retrieved |
| --- | --- | --- |
| #1 Population | (MH "Myocardial Ischemia+") OR (MH "Coronary Artery Bypass+") OR (MH "Myocardial Revascularization+") OR (MH "Heart Failure+") OR myocard* N3 isch?mi* OR isch?mi* N3 heart OR coronary OR myocard* N3 infarct* OR heart N3 infarct* OR angina OR heart N3 attack OR myocard* OR cardiac* OR cabg OR ptca OR (stent* N3 (heart OR cardiac*)) OR (stent* N3 (heart OR cardiac*)) OR heart N3 failure | 359,810 |
| #2 Concept | increas* N3 (enlist* OR enrol* OR attend* OR engage* OR "sign up" OR "take up" OR uptake OR comply* OR complian*) OR increas* N3 participat* OR motivat* N3 interv* OR increas* AND (MH "Patient Compliance+") | 32,441 |
| #3 Context | (MH "Rehabilitation Centers+") OR (MH "Therapeutic Exercise+") OR (MH "Exercise+") OR (MH "Physical Fitness+") OR (MH "Patient Education+") OR (MH "Self Care+") OR (MH "Ambulatory Care") OR (MH "Counseling+") OR (MH "Health Education+") OR MH rehabilitation, Cardiac OR "Cardiac rehabilitation" OR physical* N3 (fit* OR train* OR therap* OR activit*) OR train* N3 (strength* OR aerobic OR exercis*) OR (exercis* OR fitness) N3 (treatment OR intervent* OR | 1,339,976 |
|  | program*) OR patient* N3 educat* OR (lifestyle OR life-style) N3 (intervent* OR program* OR treatment*) OR councel?ing OR stress N3 manage* OR manage* N3 (anxiety OR depres*) OR ((behavior* OR behaviour*) N4 (modify OR modificat* OR therap* OR change)) OR goal N3 setting OR (nutrition OR diet OR health) N3 educat* OR heart manual OR ("cardiac rehabilitation" OR intervent* OR program* ) |  |
|  | #1 AND #2 AND #3 | 976 |
| Limited to studies published between 2000 and the present<br>Limited to adults > 18 years<br>Limited to language: English, Danish, Swedish, Norwegian and German |  |  |

### Source of evidence selection

Following the search, all identified records will be collated and uploaded into the software management program Covidence. After removing duplicates, titles and abstracts will be screened by two independent reviewers for assessment against the inclusion criteria for the review. Potentially relevant papers will be retrieved in full text and will be assessed in detail against the inclusion criteria by two independent reviewers. Reasons for exclusion of full-text papers that do not meet the inclusion criteria will be recorded and reported. Any disagreements that arise between the reviewers at any stage of the selection process will be resolved through discussion or with a third reviewer. The results of the search will be reported in full in the final scoping review and presented in a Preferred Reporting Items for Systematic Reviews and Meta-analyses for Scoping Reviews (PRISMA-ScR) flow diagram [36]

### Data extraction

Data will be extracted from papers included in the scoping review by two independent reviewers using a data extraction tool developed by the reviewers. The data extracted will include specific details about adults included in studies on face-to-face interventions to encourage enrolment in CR. For clinical trials, observational studies, and descriptive studies study characteristics, i.e., design, purpose, and main outcomes, as well as intervention characteristics, i.e., intervention components and strategies. Additionally, key findings relevant to the review question will be extracted. For qualitative studies also experiences, barriers and facilitators will be extracted. Draft extraction tools are provided (see Table II and Table III). The draft data extraction tools will be modified and revised as necessary during the process of extracting data from each included paper. Modifications will be detailed in the full scoping review. Any disagreements that arise between the reviewers will be resolved through discussion or with a third reviewer. The authors of the included papers will be contacted to request missing or additional data, where required.

### Presentation of the results

The process of identification, selection, and exclusion of full text studies will be visualized in a PRISMA flow diagram. We will present the findings in a narrative form with a description of the face-to-face interventions and their relation to the review objective and questions. A summary of the extracted data, key concepts, and recommendations will be provided, and an effort will be made to identify knowledge gaps. A description of common themes and differences across the interventions will be provided. Key findings will be mapped and presented in diagrammatic or tabular form as we deem appropriate considering the nature of the findings. The tables for data presentation will be developed in a refined version based on the data charting forms presented in Table II and III.

**Table II:** Data extraction instrument for clinical trials and observational / descriptive studies

| Reference | Study characteristics |  |  |  |  |
| --- | --- | --- | --- | --- | --- |
| Author,<br>year,<br>country | Study purpose and study design | Setting and length of intervention | Intervention deliverer | Population / sample size, mean age, admission diagnosis | Main outcome(s) and the effectiveness |
|  | Intervention characteristics |  |  | Findings |  |
|  | Intervention format and components | Theory or model used | Patients' experiences taken into account | Strategies used | Main findings |

**Table III:** Data extraction instrument for qualitative studies.

| Reference | Study characteristics |  |  |  |
| --- | --- | --- | --- | --- |
| Author, Year, country | Study purpose and study design | Theoretical perspective | Intervention deliverer | Population / sample size, mean age, admission diagnosis |
|  | Study characteristics | Results described by authors related to enrolment |  |  |
|  | Intervention characteristics | Experiences | Facilitators | Barriers |

## Data Availability

The study is a scoping review protocol and the manuscript does not refer to any data

## ETHICS AND DISSSEMINATION

Since the scoping review methodology aims at synthetizing information from publicly available publications, this study does not require ethical approval. In terms of dissemination activities, an article reporting the results of the scoping review will be submitted for publication to a scientific journal and presented at relevant conferences. We expect the results of the scoping review will provide a comprehensive overview of the evidence base of face-to-face interventions to encourage enrolment in CR and to highlight areas where evidence is missing. The results may be of interest for health professionals interested in planning and delivering evidence based and effective interventions to encourage participation in CR. For this reason, the results will be also disseminated as part of future workshops with professionals involved in communicating with patients about CR.

## Authors’ Contributions

BRA drafted the protocol and was the main writer of the manuscript and responsible for the initial design of this study. BST and SJH contributed to the conceptualization of the review design, specifically in establishing the inclusion and exclusion criteria. BRA executed the draft search strategy. BST contributed to developing the extraction criteria, provided feedback on the background section, and revised the manuscript for content and clarity. All authors read, edited, and approved the final manuscript prior to submission.

## Acknowledgements

We thank Research Secretary Line Jensen, Department of Research, Horsens Hospital, Denmark for proofreading and Librarian, Karin Friis Velbæk, Medical Library, Viborg Regional Hospital for assisting in producing the literature search plan.

## REFERENCES

1. Roth GA, Johnson C, Abajobir A, et al. Global, Regional, and National Burden of Cardiovascular Diseases for 10 Causes, 1990 to 2015. Journal of the American College of Cardiology 2017;70(1):1–25.

2. Levy D, Kenchaiah S, Larson MG, et al. Long-term trends in the incidence of and survival with heart failure. N Engl J Med 2002;347(18):1397–402.

3. Norton C, Georgiopoulou VV, Kalogeropoulos AP, et al. Epidemiology and cost of advanced heart failure. Prog Cardiovasc Dis 2011;54(2):78–85.

4. Le J, Dorstyn DS, Mpofu E, et al. Health-related quality of life in coronary heart disease: a systematic review and meta-analysis mapped against the International Classification of Functioning, Disability and Health. Qual Life Res 2018;27(10):2491–503.

5. Anderson L, Oldridge N, Thompson DR, et al. Exercise-Based Cardiac Rehabilitation for Coronary Heart Disease: Cochrane Systematic Review and Meta-Analysis. Journal of the American College of Cardiology 2016;67(1):1–12.

6. Long L, Mordi IR, Bridges C, et al. Exercise-based cardiac rehabilitation for adults with heart failure. Cochrane Database Syst Rev 2019;1(1):Cd003331.

7. Meijer A, Conradi HJ, Bos EH, et al. Prognostic association of depression following myocardial infarction with mortality and cardiovascular events: a meta-analysis of 25 years of research. General hospital psychiatry 2011;33(3):203–16.

8. Mampuya WM. Cardiac rehabilitation past, present and future: an overview. Cardiovascular diagnosis and therapy 2012;2(1):38–49.

9. Samayoa L, Grace SL, Gravely S, et al. Sex differences in cardiac rehabilitation enrollment: a meta-analysis. The Canadian journal of cardiology 2014;30(7):793–800.

10. Grace SL, Turk-Adawi KI, Contractor A, et al.Cardiac Rehabilitation Delivery Model for Low-Resource Settings: An International Council of Cardiovascular Prevention and Rehabilitation Consensus Statement. Progress in cardiovascular diseases 2016;59(3):303–22.

11. Balady GJ, Ades PA, Bittner VA, et al. Referral, enrollment, and delivery of cardiac rehabilitation/secondary prevention programs at clinical centers and beyond: a presidential advisory from the American Heart Association. Circulation 2011;124(25):2951–60.

12. Piepoli MF, Hoes AW, Agewall S, et al. 2016 European Guidelines on cardiovascular disease prevention in clinical practice: The Sixth Joint Task Force of the European Society of Cardiology and Other Societies on Cardiovascular Disease Prevention in Clinical Practice (constituted by representatives of 10 societies and by invited experts)Developed with the special contribution of the European Association for Cardiovascular Prevention & Rehabilitation (EACPR). European heart journal 2016;37(29):2315–81.

13. Simony CP, Dreyer P, Pedersen BD, et al. Empowered to gain a new foothold in life--A study of the meaning of participating in cardiac rehabilitation to patients afflicted by a minor heart attack. International journal of qualitative studies on health and well-being 2015;10:28717.

14. Clark AM, King-Shier KM, Spaling MA, et al. Factors influencing participation in cardiac rehabilitation programmes after referral and initial attendance: qualitative systematic review and meta-synthesis. Clinical rehabilitation 2013;27(10):948–59.

15. Dansk Hjerterehabiliteringsdatabase (DHRD). Dokumentalistrapport, version 2.0, November 2018.

16. Dalal HM, Doherty P, Taylor RS. Cardiac rehabilitation. Bmj 2015;351:h5000.

17. Doherty PJ, Harrison AS. The National Audit of Cardiac Rehabilitation: Quality and Outcomes Report 2018. 2018.

18. Resurrección DM, Moreno-Peral P, Gómez-Herranz M, et al. Factors associated with non-participation in and dropout from cardiac rehabilitation programmes: a systematic review of prospective cohort studies. European Journal of Cardiovascular Nursing 2019;18(1):38–47.

19. Ruano-Ravina A, Pena-Gil C, Abu-Assi E, et al. Participation and adherence to cardiac rehabilitation programs. A systematic review. International journal of cardiology 2016;223:436–43.

20. Gravely-Witte S, Leung YW, Nariani R, et al. Effects of cardiac rehabilitation referral strategies on referral and enrollment rates. Nature reviewsCardiology 2010;7(2):87–96.

21. Santiago de Araujo Pio C, Chaves GS, Davies P, et al. Interventions to promote patient utilisation of cardiac rehabilitation. The Cochrane database of systematic reviews 2019;2:CD007131.

22. Scott LB, Gravely S, Sexton TR, et al. Examining the effect of a patient navigation intervention on outpatient cardiac rehabilitation awareness and enrollment. J Cardiopulm Rehabil Prev 2013;33(5):281–91.

23. Carroll DL, Rankin SH, Cooper BA. The effects of a collaborative peer advisor/advanced practice nurse intervention: cardiac rehabilitation participation and rehospitalization in older adults after a cardiac event. J Cardiovasc Nurs 2007;22(4):313–9.

24. Ali-Faisal SF, Benz Scott L, Johnston L, et al. Cardiac rehabilitation referral and enrolment across an academic health sciences centre with eReferral and peer navigation: a randomised controlled pilot trial. BMJ Open 2016;6(3):e010214.

25. Rouleau CR, King-Shier KM, Tomfohr-Madsen LM, et al. The evaluation of a brief motivational intervention to promote intention to participate in cardiac rehabilitation: A randomized controlled trial. Patient education and counseling 2018;101(11):1914–23.

26. Cossette S, Frasure-Smith N, Dupuis J, et al. Randomized controlled trial of tailored nursing interventions to improve cardiac rehabilitation enrollment. Nurs Res 2012;61(2):111–20.

27. Dankner R, Drory Y, Geulayov G, et al. A controlled intervention to increase participation in cardiac rehabilitation. Eur J Prev Cardiol 2015;22(9):1121–8.

28. Mosleh SM, Bond CM, Lee AJ, et al. Effectiveness of theory-based invitations to improve attendance at cardiac rehabilitation: a randomized controlled trial. Eur J Cardiovasc Nurs 2014;13(3):201–10.

29. Desveaux L, Saragosa M, Russell K, et al. How and why a multifaceted intervention to improve adherence post-MI worked for some (and could work better for others): an outcome-driven qualitative process evaluation. BMJ Open 2020;10(9):e036750.

30. Todres L, Galvin K, Dahlberg K. Lifeworld-led healthcare: revisiting a humanising philosophy that integrates emerging trends. Medicine, health care and philosophy 2007;10(1):53–63.

31. Galvin KT, Todres L. Kinds of well-being: A conceptual framework that provides direction for caring. International journal of qualitative studies on health and well-being 2011;6(4):10.3402/qhw.v6i4.10362. Epub 2011 Dec 9.

32. Dahlberg K, Todres L, Galvin K. Lifeworld-led healthcare is more than patient-led care: An existential view of well-being. Medicine, Health Care and Philosophy 2009;12(3):265–71.

33. Karmali KN, Davies P, Taylor F, et al. Promoting patient uptake and adherence in cardiac rehabilitation. The Cochrane database of systematic reviews 2014;(6):CD007131. doi(6):CD007131.

34. Uhrenfeldt L, Sørensen EE, Bahnsen IB, et al. The centrality of the nurse-patient relationship: a Scandinavian perspective. Journal of Clinical Nursing 2018.

35. Aromataris E, Munn Z. JBI manual for evidence synthesis. Adelaide: The Joanna Briggs Institute 2020 [cited Okt 12, 2020].

36. Tricco AC, Lillie E, Zarin W, et al. PRISMA extension for scoping reviews (PRISMA-ScR): checklist and explanation. Annals of internal medicine 2018;169(7):467–73.

